# Plug-and-play microphones for recording speech and voice with smart devices

**DOI:** 10.1101/2023.09.30.23296391

**Authors:** Gustavo Noffs, Matthew Cobler-Lichter, Thushara Perera, Scott C. Kolbe, Helmut Butzkueven, Frederique M. C. Boonstra, Anneke van der Walt, Adam P. Vogel

## Abstract

**INTRODUCTION:** Smart devices are widely available and capable of quickly recording and uploading speech segments for health-related analysis. The switch from laboratory recordings with professional-grade microphone set ups to remote, smart device-based recordings offers immense potential for the scalability of voice assessment. Yet, a growing body of literature points to a wide heterogeneity among acoustic metrics for their robustness to variation in recording devices. The addition of consumer-grade plug-and-play microphones has been proposed as a possible solution. The aim of our study was to assess if the addition of consumer-grade plug-and-play microphones increases the acoustic measurement agreement between ultra-portable devices and a reference microphone.

**METHODS:** Speech was simultaneously recorded by a reference high-quality microphone commonly used in research, and by two configurations with plug-and-play microphones. Twelve speech-acoustic features were calculated using recordings from each microphone to determine the agreement intervals in measurements between microphones. Agreement intervals were then compared to expected deviations in speech in various neurological conditions. Additionally, each microphone’s response to speech and to silence were characterized through acoustic analysis to explore possible reasons for differences in acoustic measurements between microphones. Lastly, the statistical differentiation of two groups, neurotypical and people with Multiple Sclerosis, using metrics from each tested microphone was compared to that of the reference microphone.

**RESULTS:** The two consumer-grade plug-and-play microphones favoured high frequencies (mean centre of gravity difference ≥ +175.3Hz) and recorded more noise (mean difference in signal-to-noise ≤ -4.2dB) when compared to the reference microphone. Between consumer-grade microphones, differences in relative noise were closely related to distance between the microphone and the speaker’s mouth. Agreement intervals between the reference and consumer-grade microphones remained under disease-expected deviations only for fundamental frequency (*f0*, agreement interval ≤0.06Hz), *f0* instability (*f0* CoV, agreement interval ≤0.05%) and for tracking of second formant movement (agreement interval ≤1.4Hz/millisecond). Agreement between microphones was poor for other metrics, particularly for fine timing metrics (mean pause length and pause length variability for various tasks). The statistical difference between the two groups of speakers was smaller with the plug-and-play than with the reference microphone.

**CONCLUSION:** Measurement of *f0* and F2 slope were robust to variation in recording equipment while other acoustic metrics were not. Thus, the tested plug-and-play microphones should not be used interchangeably with professional-grade microphones for speech analysis. Plug-and-play microphones may assist in equipment standardization within speech studies, including remote or self-recording, possibly with small loss in accuracy and statistical power as observed in the current study.

## INTRODUCTION

Acoustic analysis forms one of the pillars in the assessment of voice quality (1). In addition, speech analysis can provide useful insights into the diagnosis and progression of various neurological conditions. Some acoustic metrics of speech are related to the severity of neurological impairment or may assist in distinguishing between healthy subjects (HS) and people with diseases such as depression (2), Multiple Sclerosis (3, 4), Parkinson’s disease (5) and others (6–10). Moreover, the acquisition of speech recordings for health-related analysis has become increasingly accessible through the use of mobile devices, both in research and clinical settings, offering potential scalability and continued care through telehealth when access becomes restricted (11).

The reliability of speech metrics from mobile device recordings remains disputed. Several studies found that some speech measurements related to mean spectral and cepstral frequencies (e.g., mean fundamental frequency, *F0*) were comparable between professional-level and consumer-grade recording equipment while measures of perturbation (e.g., shimmer) presented much more variation in agreement between recording equipment (12–21). To that effect, Daudet et al. (2017) (6) argued that the in-built microphones of mobile devices were insufficient to filter out noise and that a high-quality microphone coupled to a mobile device was necessary to achieve the high accuracy for detecting traumatic brain injury in out-of-lab conditions. Conversely, Rusz et al. (2018) (15) found that speech metrics from smartphone recordings were almost as sensitive as the professional grade microphone in capturing (previously studied) key abnormalities in speech of people at high risk of developing or already diagnosed with Parkinson’s disease.

Various factors may account for the variability in speech metrics’ results. In contrast to research experiments, clinical practice and self-recording often introduce uncontrolled variables such as background noise, variation in the distance and direction between microphone and sound source, and differences in microphones’ properties. To quantify the influence of some of these variables on measurements’ results, Titze and Winholtz varied the distance between professional-grade microphones and a synthesized voice source from 4cm to 30cm to 1m. They observed that the recorded amount of false signal perturbation (i.e., noise or aperiodicity) progressively increased by almost one order of magnitude at each distance-step, for both frequency and amplitude. Similarly, the use of consumer-grade microphones resulted in greater false perturbations and less responsiveness to true perturbations, particularly to increased aperiodicity, when compared to professional-grade microphones at a fixed distance. These findings were associated with the microphone’s sensitivity at 200Hz, which is close to the fundamental frequency of human voice (22). A greater sensitivity to frequencies occupied by voice improves the signal-to-noise ratio (SNR). The same logic applies for the positioning of microphones where proximity to sound source results in a greater target signal magnitude while keeping constant the recorded amplitude of background noise, i.e., higher SNR and consequently smaller relative noise. Relative noise, in turn, would interfere with various acoustic measurements related to variation and periodicity (12, 23) (e.g., *f0* standard deviation, harmonics-to-noise ratio, cepstral peak prominence - CPP) which are commonly used for voice analysis.

Microphones’ sensitivity varies across frequencies. The distribution of sensitivity along the frequency spectrum is referred to as frequency response, which often influences the recorded signal in a predicable manner (i.e., systematic deviations). The importance of accounting for the microphone’s frequency response was highlighted in an experiment by Parsa et al. (2001) (24). The authors used loudspeaker playback of human voices to compare four professional-grade microphones and found that systematic divergencies in microphones’ frequency responses accounted for small distortions in acoustic analysis, but which were sufficient to decrease the accuracy in discriminating between normal and pathological voices. Because of the largely systematic nature of the differences, a post-recording mathematical correction accounting for the microphone’s frequency response improved the accuracy for discriminating between healthy and pathological voices. In a recent study, Awan et al compared playback speech recorded by four modern smartphones, which had similar frequency responses, and a reference microphone and observed a correlational equivalence in measured CPP but a large device-effect on measured spectral tilt (25). In a similar study testing consumer and professional-grade head-worn microphones, this time with different frequency response curves, Awan et al highlighted that differences in measured CPP were not correlated to frequency response or to microphone’s sensitivity around 100-200Hz (26) in contrast to earlier assumptions (22). Thus, frequency response alone seems insufficient to predict the accuracy of perturbation measurements.

While recording variables responsible for variation in measured speech acoustics (e.g., distance to microphone, frequency response curve) remain under investigation, a pragmatic approach may benefit the scalability in speech research. In that sense, agreement intervals, random and systematic errors are reliability statistics may be compared to accuracy requirements to inform the appropriateness of a certain equipment. With exception of Jannets et al (16), studies did not report random error or agreement intervals between microphones, and none directly compared random measurement error to expected disease or treatment effects, i.e., clinical significance of measurement error. In the current pilot experiment, we aimed to test if from the addition of portable and affordable consumer-grade microphones increased measurement agreement against a typical reference equipment. We tested two mount-on microphones for mobile devices, which could be used in conjunction with “bring your own” iOS or Android device in studies. We extracted intensity-related, frequency-related, and periodicity metrics from simultaneous recordings and calculated differences, agreement intervals, and correlation coefficients between microphones. Based on previous findings, we hypothesize that agreement intervals between microphone set ups would be smaller than the expected clinical effect (thus acceptable) for *f0* and for intensity-related timing measurements but not for metrics of perturbation-periodicity or other frequency-related measurements (16–18, 25, 26). Finally, we hypothesize that despite possible differences in measurements, the consistent use any of the tested microphones would yield a similar differentiation between a group of dysarthric (mild and sub-clinical) and non-dysarthric speakers.

## METHODS

The study was reviewed and approved by the Melbourne Health Ethics Committee (approval number 2015.069). All participants were informed of the purpose of the study as well as their right to withdraw at any point, and signed consent was obtained.

### Materials

We selected a high-quality configuration previously used in speech research as the reference microphone, consisting of an AKG C520 head-worn cardioid condenser microphone coupled to a Roland Duo Capture EX USB Audio Interface and connected to a laptop. Characteristics of this microphone include a sensitivity of 5mV/Pa, a near flat tonal frequency response, a condenser transducer type, cardioid pattern, and mount positioning close to sound source, which are considered adequate for recording speech for acoustic analysis according to various reference authors (22, 26–28). Consumer grade microphones included three configurations of the 6^th^ generation iPod Touch: (1) the in-built iPod Touch microphone (in-built); (2) Rode IXY-L mobile-mount cardioid condenser microphone (directional, sensitivity of 8.5mV/Pa); and (3) Sennheiser ClipMic Digital lapel omnidirectional condenser microphone (lapel, sensitivity of 5mV/Pa).

### Participants

In addition to people with normal voice and speech, we purposefully recruited people with dysarthria to increase the spread of each acoustic metric, which assists in gauging the dependency between inter-microphone differences (bias and random error) and the magnitude of the measurement. Checking for such dependency is one of the steps in measuring agreement intervals through the elected method further described under statistical analysis. In total, eighteen people (10 females) aged between 18 and 59 years participated in the study and were included in the analysis. Inclusion of people with dysarthria was achieved through five participants with Multiple Sclerosis (pwMS), which included two people with mild dysarthria and three people with clinically normal speech, all of whom were included in the analysis.

For simplicity, we excluded recordings of sustained vowel from one female HS who produced intermittent but frequent vocal fry, which caused extraneous variation (halving) in *f0* measurements. Additionally, recordings of two people with MS were summarily terminated by the device connected to the directional microphone (corrupted data) and thus unsuitable for acoustic analysis.

### Tasks

We used four increasingly demanding speech tasks to elicit increasing complexity in acoustics. These tasks included (1) sustaining the vowel /a/ for ten seconds; (2) repeating /pa/ta/ka/ as fast as possible for ten seconds (speech diadochokinesis task, DDK); (3) reading a phonetically and linguistically balanced text, the grandfather passage; and (4) a one-minute monologue consisting of recounting a pleasant memory (free speech). These tasks are designed to test specific aspects of speech and are commonly used in voice and speech research (12, 13, 18, 29–37).

### Recording protocol

Recordings occurred simultaneously through the four sets of equipment in a hospital outpatient clinic room in a quiet corridor away from main thoroughfare and reception areas.

The design and build of a microphone determine its fixed properties (e.g., self-generated noise, sensitivity, frequency response) but also its typical positioning in relation to sound source (e.g., table-top microphone, hand-held, head-mount, lapel-clip). Thus, to emulate their use by a hypothetical consumer (patient or clinician), each microphone was positioned according to its commercial design as follows. The reference microphone was positioned at an angle of 45 degrees laterally to the sagittal plane, axially at the level of the mouth, at approximately 8cm from the participants’ mouths, and supported by a back-of-head and around-ears adjustable arch. The in-built and the directional microphones were kept on top of a table directly in front of participants, 80 cm above the floor level, and approximately 50cm away from the participants’ mouths. To ensure proper orientation of the directional microphone towards the participants’ mouths, its respective iPod was secured in place with a Joby Grip Tight Pro Mount tripod which elevated the device about 10cm from the tabletop. The lapel microphone was secured to the participants’ clothes (e.g., shirt) over the anterior thorax in a position that would not cause friction or inadvertent motion artifact, at approximately 20cm from their mouth. Those fixed microphone positions were used to record each participant. Unless otherwise specified, these exact recording configurations (microphone + iPod/laptop + positioning) will be referred to as simply ‘microphones’ (or their specific designation, e.g., ‘lapel’) from this point forward. Recording and pre-analyses procedures are further detailed in supplementary methods.

### Acoustic analyses

Acoustic analysis was performed in the open-source software Praat (38). Two groups of speech features were analysed, the first of which aimed at determining agreement between measurements taken by different microphones (main study aim). We determined the inter-microphone limits of agreement(39) separately for twelve acoustic measurements (Table 1). Methodological details on acoustic analysis, limits of agreement and statistical analysis are included in supplementary methods.

To assist in descriptively exploring variables responsible for possible low agreements between microphones, we further analysed each microphone’s responsiveness to speech and silence. We characterized microphones’ responsiveness to speech frequencies instead of pure tones. Pure tone responsiveness curves are available online for the reference and directional microphones. Responsiveness to speech accounts for possible interactions between components of the complex voice waveform as well as to voiceless transitions and silence-speech boundaries and does not require extra data collection. Frequency responsiveness was characterized by power spectral density (PSD, Figure 1) and its weighted average, centre-of-gravity (CoG). Additionally, SNR was determined for each participant’s microphone and reported as average and confidence interval per microphone. PSD, CoG, and SNR were determined over synchronized recordings of the reading passage while all frequency and periodicity metrics were derived from the sustained vowel. Lastly,

**Figure 1.**
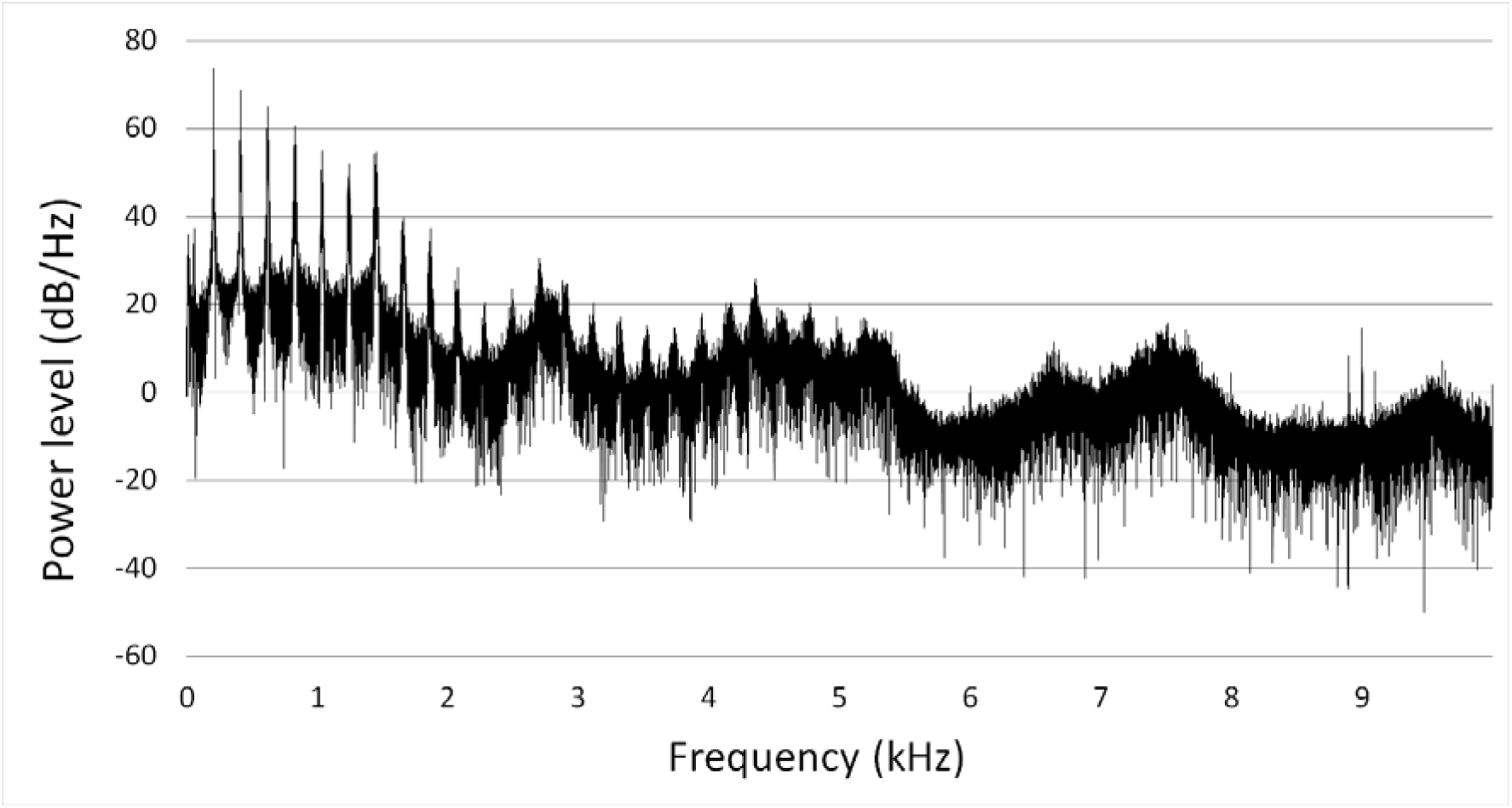
Power spectral density levels for mid-vowel /a/, participant 3 (female without MS). The first peak represents the *f0* followed by peaks of other harmonics. Frequencies above 2kHz have low power peaks whereas no distinct peaks are observable above 5kHz.

Limits of agreement and confidence intervals alone are insufficient to determine whether devices can be used interchangeably in research and clinical settings. Agreement intervals between measurements must be examined in relation to the expected differences between conditions of interest (e.g., affected people vs HS, dysarthric vs non-dysarthric). For an overview of the clinical relevance of our findings, we reproduced differences between clinical conditions found in previous studies. In general, if the coefficient of agreement for a pair of microphones is greater than the expected difference between clinical conditions, there would be insufficient reliability for using those microphones interchangeably to measure differences between conditions.

Lastly, we explored how metrics from each microphone would differentiate two groups of speakers. In a previous work (40), we have modelled an acoustic composite score to reflect general neurological disability in pwMS consisting of pause percentage in the monologue, DDK rate, and *f0* CoV in the sustained vowel. Via Spearman’s and Pearson’s correlation coefficients, we first compared rank and linear classification of speech between microphones for each component in the acoustic composite score. We also calculated the acoustic composite score separately for each microphone’s recordings and plotted the results for comparison. The neurological score of participants with MS was determined by a neurologist through the Expanded Disability Status Scale (EDSS) score (41). Given the limited number of pwMS, comparing graphs between microphones for this simulated neurological classification is merely exploratory. Finally, we report how the acoustic scores from each microphone would differentiate the groups of neurotypical and pwMS.

## RESULTS

### Microphone characteristics

The intensity of recorded signal during speech intervals (i.e., noise during silence) was greater for the in-built microphone relative to the reference microphone (intensity floor, Table 2). Higher relative noise for the in-built microphone can also be observed during phonation and in more detail on curves of power spectral density (PSD, Figure 2) where the 25dB gap in responsiveness to *f0* between reference and in-built (first marker in each curve, under 0.3kHz) disappear for frequencies commonly populated by noise (>5kHz). While the three consumer-grade microphones were proportionally more sensitive to higher frequencies in comparison to the reference microphone (higher CoG, Table 2), that difference was more associated with responsiveness to the *f0* and less with noise for both lapel and directional microphones (Table 2 and Figure 2).

**Figure 2.**
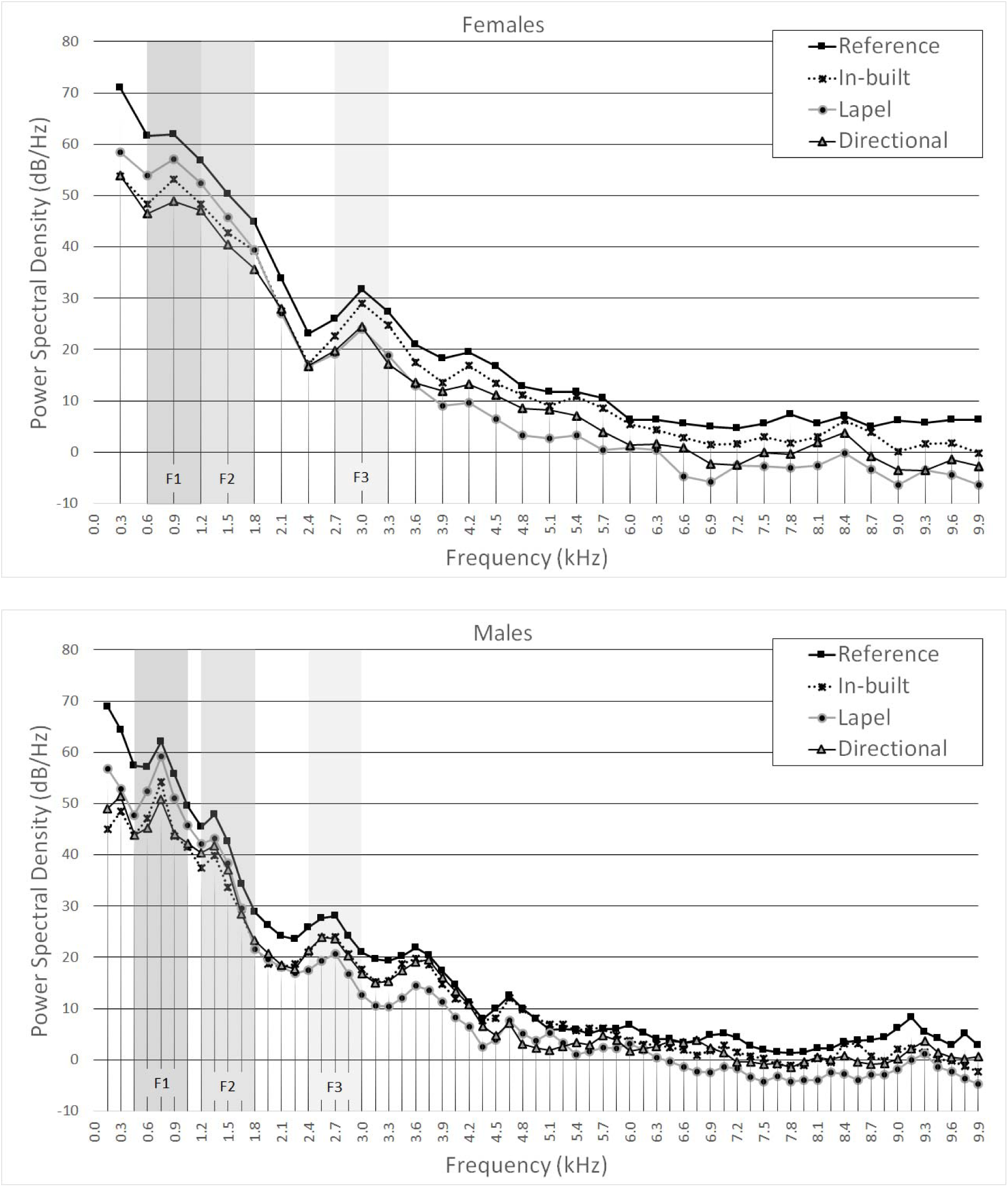
Average power spectral density (PSD) levels (re 10^-5^ Pa) for voices of males and females, by microphone. The first marker on each line represents the power in *f0* whereas shaded areas highlight sound power for formant frequencies (as determined by analysis of recordings by the reference microphone). F1, F2, F3 = first three formants.

### Agreement coefficients

Agreement between microphones was highest for *f0* measurements (Tables 2 and 3, Figure 3). Coefficients of agreement and 95%CIAs remained under 0.1Hz for mean *f0*and under 0.05% for *f0* CoV. Measurements of average formant frequency, HNR and CPP diverged 10% to 25% between microphones. All three formants’ SD presented the largest 95% CIA, up to ten times as wide as the formant’s average SD, i.e., 1000% divergence in measurements.

**Figure 3.**
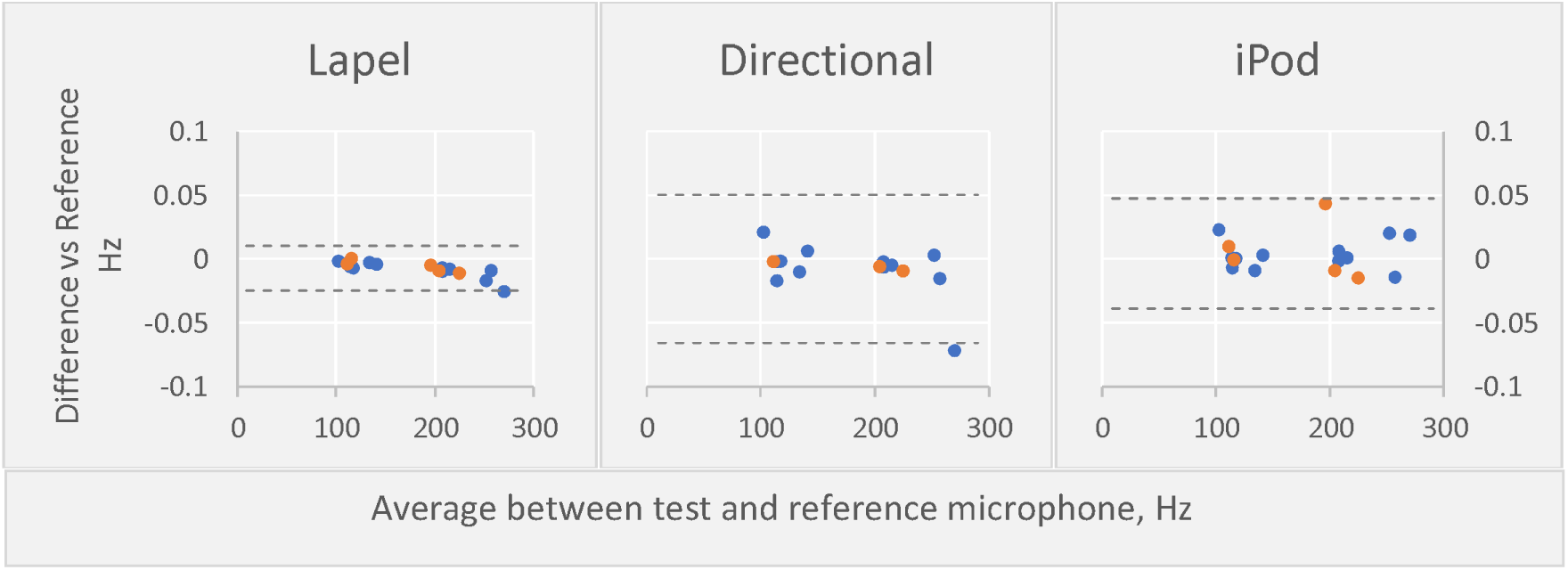
Bland-Altman plots and 95% confidence interval of agreement (dotted lines) for *f0*, showing good agreement between reference and test microphones. PwMS are shown in red. The average difference between microphones is close to zero, i.e., not systematic (small measurement bias), and differences are independent of average measurement or of presence of MS. Random disagreement is also small, highlighted by 95% CIA under 0.1% of the average measurement.

Among timing measurements (Table 3), utterance sum for the reading and free speech tasks presented the most agreement between microphones (5% to 7.5% as measured by 95% CIA) followed by pause length SD in the same tasks and pause percentage in all tasks (20% to 30%, Figure 4). Other measurements had 95% CIAs wider than 30% of the mean for that measurement.

**Figure 4.**
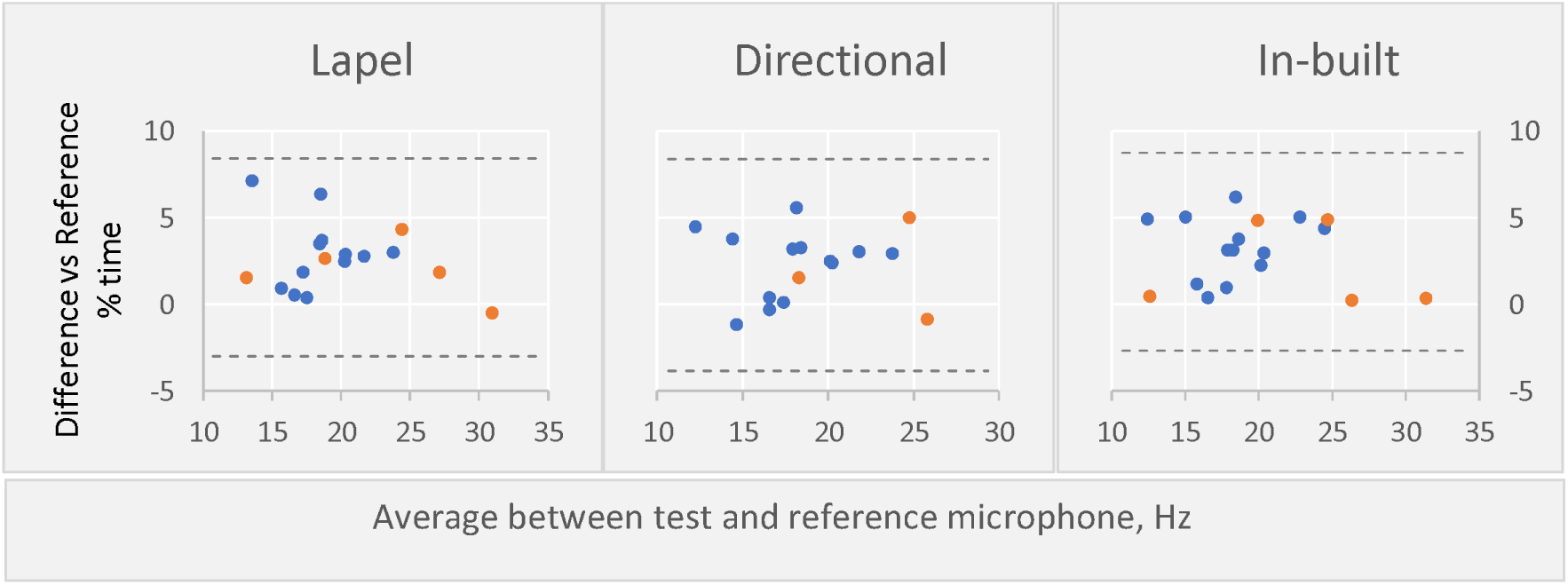
Bland-Altman plots and 95% CIA (dotted lines) for monologue pause percentage, showing poor agreement between reference and test microphones. PwMS are shown in red. All test microphones systematically overestimated pause percentage in comparison to the refence microphone (measurement bias) but differences seem independent of average measurement or of presence of MS. Non-systematic (random) disagreement is reflected in large 95% CIA, which are over 20% of the average measurement.

Limits of agreement between the directional and reference microphones for DDK mean syllable period could not be calculated as differences were not normally distributed. Similarly, raw and log transformed differences between microphones for the acoustic composite score were not normally distributed, thus not suitable for calculation of agreement intervals through the elected method.

### Classificatory comparison between microphones

Rank classification and linear values of speech acoustic metrics were compared between each pair of microphones. Both Spearman’s rank and Pearson’s linear correlation coefficients between any pair of microphones were r >0.86 (p<0.001) for pause percentage in free speech, r >0.80 (p <0.001) for mean DDK rate, r >0.99 (p <0.001) for *f0* CoV and r>0.79 (p <0.001) for the acoustic composite score.

Group comparison for the acoustic composite scores between HS and MS yielded Mann-Whitney asymptomatic significance values of p=0.015 for the reference, p=0.04 for lapel and p=0.08 for the in-built microphone. Spearman’s correlation coefficient between EDSS and the acoustic composite score was the same, r =0.84 (p=0.04), regardless of microphone considered. The scoring of speech from dysarthric and non-dysarthric subjects was also similar between microphones as visualized in Figure 5.

**Figure 5.**
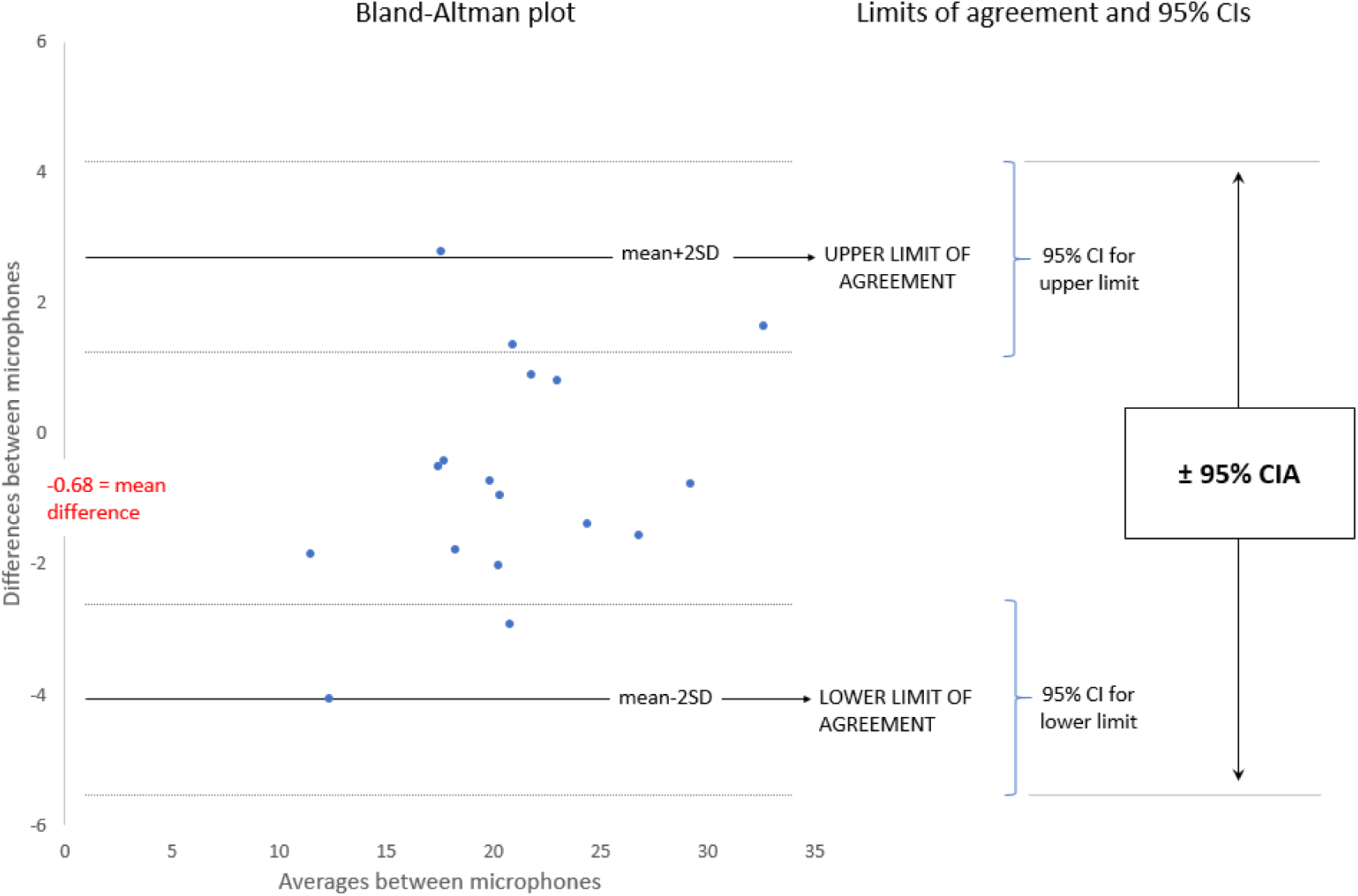
Scores form the acoustic composite score according to each microphone. HS = healthy subjects; MS = participants with Multiple Sclerosis

## DISCUSSION

We found that measurements of *f0* were robust to variation in recording device sets, which is consistent with previous findings comparing various mobile recording devices to a high-quality studio microphone in both vocally healthy adults and subjects with voice disorders (16, 17, 42, 43). In addition, F2 slope values had agreement intervals marginally smaller than clinical effect sizes. Other measurements, including all timing metrics, were largely influenced by the recording device type, which does not support our initial hypothesis. In partial contrast to previous findings (15), differentiation between neurotypical and pwMS decreased slightly with the use of the consumer-grade microphones despite the high correlation of measurements between reference and tested microphones.

Previous studies also found that jitter and shimmer, which are short-term perturbation metrics of the *f0* (of its period and intensity, respectively) were reasonably consistent across different recording devices (20, 43) and ambient noise (19). The following characteristics of the *f0* allow it to be estimated through simple methods, which might explain its robustness observed in the current and previous studies. The quasi-periodic vibration of the vocal folds (glottis) during phonation is responsible for the *f0*, which is theoretically the lowest periodic frequency in the voice waveform. Because this glottal sound is the major energy source for production of other sounds in voice, its energy relative to the total voice energy is great enough to be detected through direct analysis of the raw (unprocessed, untransformed) recorded signal. By default, the *f0* is determined by the auto-correlation method in Praat, which consists of simply making an identical copy of the signal and then calculating the correlation coefficient between original and copy each time the copy is slid forward by a chosen amount in the time domain. The predominantly periodic nature of voice means that the sound wave “repeats itself” at almost fixed intervals, at least in short time windows. Thus, sliding the copy by exactly one such “repeating interval” will yield a very high correlation coefficient between copy and original. The amount of displacement of the copy for which the highest correlation coefficient is found defines the size of the repeating interval, called fundamental period. The *f0*, in turn, refers to the number of periods that occur per time (per second, in case of Hertz). Because the auto-correlation method examines the whole waveform, it might be susceptible to inaccuracy in determination of the *f0* when other components of the waveform (i.e., formants) change frequencies within the analysis window. This occurs, for example, during production of more than one phonetic vowel within one utterance in naturalistic speech. Yet, that would not necessarily affect agreement between microphones, i.e., estimations would be “equally wrong” across microphones.

Other frequency-related measurements depend on decomposition of the recorded signal to produce estimates. The sound energy of higher frequencies is often many times smaller than that of the *f0* in quiet (conversational level) utterances. In the current experiment, tracking of F2 movement (i.e., F2 slope) was less affected by different microphones than steady state mean estimates (i.e., F2 in the sustained vowel). Except for the lapel microphone, the average bias for mean F2 was similar to that found by two other recent studies for desktop computer recordings (17) and for Android and iPhone smartphones (21), while the present bias for F3 with add-on microphones was smaller than the desktop computer but larger than the hand-held smartphones (17). Determining the location of formants, such as mean F2, depends on the prominence of the signal above noise plus nearby average spectral power . Separation between formant and harmonic frequencies determines a certain degree of distortion in the estimation of the former while the distance threshold for various degrees of distortion is influenced by spectral tilt (44). Thus, both microphone’s frequency response curve and noise levels may affect the estimation of the formant’s location. Estimates of a formant’s location might diverge between microphones more than estimates of that formant’s movement, e.g., F2 slope, if those estimates move in parallel between microphones (i.e., highly correlated measurements despite difference in absolute value).

We found that timing results, e.g., pause percentage, varied widely between recordings from different microphones despite no differences in sampling rate or time between events given that recordings were synchronous. The timing analysis method in the current experiment is based on sound intensity in the time domain. The threshold for separating silence and speech is determined in relation to the maximum sound pressure level which likely interacted with different SNRs between microphones to result in poor agreement as discussed below.

In our view, there are several microphone attributes that could have caused lack of agreement for measurement of timing and/or high frequencies. These include transient responses, i.e. the speed of microphones to respond to an abrupt sound (45). Different transient responses affect the onset time and onset slope of speech. As transient responses are very short by definition, they would affect more the measurement of precise (time-wise short) speech events, as consonant duration for example, and only discretely the measurement of long speech events such as utterance duration, which comprises the bulk of timing metrics analysed in the current study.

Secondly, microphone positioning might greatly affect speech measurements. Increasing the distance between sound source and microphone makes the signal more susceptible to non-free field influences (e.g. reflection and refraction of sound in out-of-lab environments) (46), which inflates measurements related to noise and perturbation or distortion (22). As expected, the decrement in SNR was proportional to the distance between microphone and mouth (source). The same decrement could be observed for clinical measurements that involve ratios between periodicity and noise, namely HNR, CPP and CPPS. The consequence of decreased SNR for timing analyses is briefly discussed below. The angle of placement in relation to the sound source can also influence measurements to a smaller degree (22) while the relation between the microphone and body parts can cause frequency-dependent distortions (47).

The third possible cause for disagreement between microphones concerns frequency responsiveness. In this study, the reference microphone’s response favoured low frequencies in comparison to the other microphones, as shown by PSD curves and the lower CoG. A microphone’s responsiveness to frequencies that naturally have higher speech energy density, particularly the *f0* in vowels, will largely influence the overall measured sound intensity. This overall intensity is the basis for determining the silence-speech threshold used in timing analyses. However, the onset and offset of syllables are not necessarily populated by low frequency energy dense speech, which creates a mismatch between the automatically determined threshold and the actual amplitude of onset/offset. Thus, optimizing the silence-speech threshold for the reference microphone causes it to be sub-optimal for another microphone and vice-versa. Frequency responsiveness further varies in relation to distance to sound source depending on the polar pattern of microphones. A noticeable boost in low frequencies is observed for cardioid microphones (e.g. the reference and the directional) when that distance is shortened, known as proximity effect (48).

Fourth, differences in relative noise level likely have also influenced timing measurements. Microphones vary in self-generated noise (45), electrical insulation and filtering of background noise. The polar pattern of microphones (directionality) also affects the amount of background noise captured (49). In our experiment, after discounting the theoretical loss in signal intensity due to the distance between microphone and mouth (source), a residual decrease in SNR of about 4dB remained for the three consumer-grade microphones relative to the reference microphone. That could be due to both reduced microphone sensitivity and increased recorded noise. The effect of SNR on HNR, CPP and CPPS was discussed above. In the current experiment, decrease in SNR means a reduced difference between speech intensity and “silence” intensity (i.e., background noise). That difference between intensities is the window for silence/speech detection. A smaller SNR means greater chance of misclassifying speech or silence for analysis based on intensity. Thus, even if the silence-speech threshold (timing analysis) was optimized individually for each microphone, its accuracy would be worse for the three consumer-grade microphones, or any microphone in conditions of excessive noise.

Those four interrelated differences were present between the tested microphone settings in the current experiment and might explain, at least in part, the lack of agreement between them. Importantly, these discussed differences may be compounded by other commonly unaccounted variables in clinical settings, such as wide variations in recording environment, bodily movements and differences in algorithms (software) used for analyses, which introduce multiple and interrelated confounders when comparing acoustic results from multiple recording protocols (50).

Limitations of the current study include low speech sample size, which increased the probability of non-normal distribution of measurements and large confidence intervals. We contacted the manufacturer (Apple Inc) asking whether the same in-built microphone sensor was present across other mobile devices (e.g., iPhones, iPads) but are yet to receive a response. Thus, although other mobile devices may produce similar agreement limits due to inherit constraints in size and power consumption, the specific agreement intervals and biases reported here relate to only the tested microphones and cannot be generalized.

In our opinion, speech research must be designed to mitigate the effect of common confounders related to hardware use. The only immediate and certain way to eliminate confounders is to standardize the speech assessment protocol within each study, which has been advocated for many years (51, 52). That includes the use of the same equipment between participants and across time in a study, fixing the distance between microphone and participant, and avoidance of noise among others. The equipment standardization between studies may be less practical in the short-term, and impossible across time given the rapid changes in technology. Accordingly, there is a trade-off between the number of confounders that are controlled in this way and the flexibility in data acquisition, which translates into a trade-off between precision and clinical scalability. Another concurrent path relates to the development of robust methods of signal analyses, which include numerous recent efforts showing variable progress. The ideal outcome is to increase robustness without compromising validity, precision, and accuracy. CPPS, for example, could be considered such an improvement over the original CPP for the estimation of dysphonia (53). In the current experiment, we observed an increased stability in measurement between different microphones for CPPS in comparison to CPP at the cost of a smaller average measurement, which is consistent with previous results (16, 19). A third path for optimizing reproducibility could be the use of combined measurements to gauge the desired outcome. Examples include principal component analysis, machine learning models or multiple regression models, such as the acoustic composite score used in the current study. Composite scores mitigate, in general, the effect of random error of its components once errors in opposite directions cancel one another. Such stabilizing effects might, however, also decrease the responsiveness of composite scores, which must be tested and weighted against its gain in reproducibility. The multiplicity of measurements used as predictors is not confined to their instantaneous qualities but could also include temporal patterns (in repeated measurement conditions). In general, the main requirements for such models are computational power (increasingly available and inexpensive) and large datasets. To produce high quality research, we must consider the variations in signal acquisition highlighted in this study and develop strategies to overcome these where possible.

## CONCLUSION

Measurement of *f0* of speech and F2 slope were robust to variation in recording equipment while other acoustic metrics were not. Most of the gain in signal-to-noise ratio for plug-and-play microphones could be explained by the microphone’s proximity to the source (i.e., to the speaker) which, however, was not proportional to the agreement in acoustic metrics when compared to the reference microphone. Metrics from plug-and-play microphones were highly correlated to the reference microphone but resulted in loss of statistical power when differentiating between neurotypical and people with MS. While we may conclude that the standardization of recording equipment is necessary for the reliable comparison of most acoustic features in repeated measure (within participants) or between participants, more research is needed to find the optimal solution.

### Statement of Ethics

This study was approved by the Melbourne Health Ethics Committee (approval number 2015.069) and was conducted in accordance with the Declaration of Helsinki. Signed consent was obtained from all study participants and they were informed of their right to withdraw participation at any point.

### Funding Sources

This work was funded by National Health and Medical Research Council of Australia, Grant 1085461, which paid for data collection costs (equipment, room, storage) as well as research stipends for Frederique M.C. Boonstra and Gustavo Noffs.

### Author Contributions

Gustavo Noffs, Anneke van der Walt, and Adam P. Vogel conceptualized and designed the study. Gustavo Noffs and Matthew Cobler-Lichter collected the data. Gustavo Noffs and Matthew Cobler-Lichter performed statistical analysis. Gustavo Noffs created the figures. Gustavo Noffs and Matthew Cobler-Lichter prepared the first draft of the manuscript. Gustavo Noffs, Matthew Cobler-Lichter, Thushara Perera, Scott C. Kolbe, Helmut Butzkueven, Frederique M. C. Boonstra, Anneke van der Walt, and Adam P. Vogel made substantial revisions to the article and gave approval of the final manuscript.

### Data Availability Statement

Data will not be made public for ethical safeguard of participants as it may be identifiable. The authors agree to share the study protocol and further details of statistical methods upon request. The experiment was not pre-registered.

## Data Availability

All available data produced in the present work are contained in the manuscript

## Tables’ legends

Table 1. Summary list of speech measurements tested for agreement.

Table 2. Comparative response characteristics of microphones for the same sound stimulus. CI = confidence interval; dB = decibel; Hz = Hertz. ^a^ Paired t-test. ^b^ Near-normal distribution. ^c^ Wilcoxon rank test.

Table 3. Bias and 95% confidence interval of agreement between reference and tested microphones for frequency-dependent measurements. CIA = 95% confidence interval of agreement; CPP(S) = cepstral peak prominence (smoothed); dB = decibel; *f0* = fundamental frequency; F(1–3) = formants; HNR = harmonics-to-noise ratio; Hz = Hertz; ms = milliseconds.

Table 4. Bias and 95% confidence interval of agreement between reference and tested microphones for timing measurements. CIA = 95% confidence interval of agreement; ms = milliseconds; s = seconds; syl = syllables.

