## Supplementary Materials for "Plug-and-play microphones for recording speech and voice with smart devices"

### Supplementary material

Recording

Audacity was used to record all speech by the reference microphone and the Rode Rec app was used to record all speech by each of the mobile devices. Before recording, digital filters and enhancers were turned off on the app. Recording parameters were kept constant for the entire experiment, including sampling rate of 44.1kHz, 16-bit quantization and constant gain (tested and set before the start of recording for each participant).

At the beginning of each recording session a pure-frequency beep (500Hz, 500ms) was played from an independent device, more than 1 meter away and equidistant from all devices, to serve as a reference time-point for later synchronization of recording files. The four microphones simultaneously recorded each participant, who produced each speech task only once. The door of the room was kept closed and recordings were stopped, discarded, and re-initiated in case of interruptions (i.e., loudspeaker announcement, knocking on the door).

Participants’ positioning in the room was also standardized. Participants were recorded while sitting, facing the center of the room and at 2 meters distance from the wall in front of them. For descriptive purposes only (rather than for room acoustic measurements), the room was rectangular and measured 3x5m, 3m high ceiling, with solid brick walls, carpeted floor and furnished with office and research equipment.

#### Pre-analysis processing

Each microphone recorded one continuous audio file per participant, including all tasks. To ensure that subsequent analyses would be carried over the same speech excerpt (i.e., synchronization), the beginning of each file was set to match the starting time-point of the initial reference beep. To ensure that time between sound-events were not significantly affected by differences in microphones’ transient response or by possible mechanical-to-digital conversion delays (e.g., due to differences in CPU processing speed while recording), the interval between the start of the beep and the start of the sustained vowel was compared between microphones. Additionally, the same beep-vowel interval was determined manually twice in each recording of two participants for all microphones (intra-screener error). The average difference between microphones was similar to the intra-screener error, on average of 10 samples (approximately 0.2ms). Thus, differences in time between speech-events between microphones were too small for detection through manual screening and could be ignored.

Next, the beginning and end of each speech task, i.e., time-boundaries, were determined manually for the audio file recorded by the Reference microphone. The maximum intra-screener error for single time-boundaries was 8 audio-samples (approximately 0.2ms) and was estimated on twenty randomly selected time-boundaries, which were determined a second time after all time-boundaries had been determined once.

The originally determined time-boundaries for each task were then applied to the other three files (in-built, directional and lapel). This process was repeated separately for each participant. Files were divided so that each post-processing file contained only one task from one microphone and from one participant, resulting in 288 speech files (18 participants x 4 tasks x 4 microphones). The files were equal in starting and finishing time across the four recording devices for each participant.

To emulate procedures applied in previous research for the analysis of specific speech features, the vowel sound task (1) was trimmed for the middle three seconds^1^ while the DDK files were trimmed to include exactly two seconds of that task after the first train of syllables (i.e. after the first ‘pataka’)^2, 3^.

##### Microphone responsiveness to speech and silence

Frequency responsiveness was characterized by power spectral density (PSD). PSD here refers to power level (in dB/Hz, relative 2x10^-5^ Pa) of the signal (i.e., speech) rather than referring to the absolute power (in Watts) of the sound source (participant). In short, PSD quantifies how much “each frequency” is contributing to the total sound power. It also allows comparison of power between different frequencies (e.g., between harmonic and noise-related frequencies) and offers a visual representation of the components of the complex waveform (figure 1). Because our aim was to compare microphones rather than compare speakers’ outputs (i.e., speech signals), PSD was averaged across participants for each microphone and compared between microphones. In voice, peaks of sound power occur at intervals very close to the *f*0^4^ (i.e. close to harmonics). For that reason, a window size slightly longer than the highest *f*0 among all participants of the same sex was used. Males and females were computed separately. For each participant, the highest peak (i.e. maximum power) within each such frequency-window was selected. Then, the average maxima among all participants of the same sex were calculated for each frequency-window. Otherwise, averaging the whole power spectrum across participants would result in an undesirable cancelation effect between peaks and troughs once each person produces power peaks at different frequencies. For visualization of differences between microphones, the average maxima for each frequency-window were plotted in a graph. Differences between microphones for a given PSD frequency-window reflect diversions in responsiveness to speech (or noise) in that frequency band. Responsiveness to frequencies of interest can then be compared (e.g., between frequencies commonly populated by the *f*0, formants and noise).

The second feature used to characterize frequency responsiveness was centre-of-gravity (CoG), also determined from the power spectrum. CoG is measured in Hz and can be understood as the power-weighted mean frequency of a sound, where the sum of power in frequencies higher than CoG equals the sum of power in frequencies lower than CoG. It was determined for each participant **on entirety of the reading task, after pre-analysis processing** (including all frequencies irrespective of peak presence), then averaged across all participants for each microphone. Average CoGs and their 95% confidence intervals were then statistically compared between microphones.

Lastly, SNR was defined as mean intensity in decibel (dB) of the vowel’s middle three seconds minus the whole file intensity floor. SNR was calculated separately for each participant-microphone file. Then the difference in SNR between the Reference and each consumer grade microphone was calculated per participant. Mean SNR differences, 95% confidence intervals (CI) and t-test p-values are reported. Because distance between microphone and participant affects the intensity of the signal of interest (speech) but not the intensity of background noise, we would expect different SNR with different distances to participant. Considering the mouth as the speech source, distance for the lapel remained at approximately 20cm/8cm=2.5 times that of the Reference, while both in-built and directional microphones rested at 50cm/8cm=6.25 times the distance between Reference and source. As intensity is inversely proportional to the square of the distance to source, and by converting to dB, distance alone would account for 10*Log10(2.5^2^) = 7.96dB loss of intensity for the lapel and 15.92dB loss of intensity for in-built and Rode. Differences between microphones in SNR corrected for distance (SNR adjusted) is also reported.

##### Determining agreement between microphones

We selected speech measurements that have been previously used in speech research related to neurological conditions and a few additional related measurements (see table 1 for a summary list). We calculated the agreement between measurements derived from recordings of different microphones **as outlined in statistical methods below**.

**With the exception of median F2 movement, all frequency and peridodicity-related metrics derived from** the VOWEL **task** which contains only continuous voicing, to mitigate possible irregular tracking of frequencies, especially formants, by the spectrum algorithm during abrupt speech transitions (voicing onset/offset and consonant-vowel). Median F2 movement^5^ **derived from a short section within the reading task**. Principles of phonetics and screening of all spectrograms directed our selection of candidate words for that measurement. The word “well” was chosen for its consistent change in F2 and for its consistent continuity in *f*0 between participants (i.e., no voice breaks, no creaky voice). **Only one “well” was produced per reading task and that was used for analysis.**

Extraction of frequency features was automated in Praat. Fundamental frequency was determined through autocorrelation using minima and maxima *f*0of 70Hz and 250Hz for men and 100Hz and 300Hz for women^6^. The analysis window was kept at 42ms and 30ms respectively, i.e., three times the size of longest wavelength, and window shift was fixed at 10ms. As per Praat guidelines and default settings, the maximum number of formants was kept at 5 with a maximum of 5500Hz for women and 5000Hz for men for formants’ detection. All other parameters were maintained at software default.

Timing measurements were also automated in Praat and separately calculated for the speech tasks DDK, reading and free speech. The detection of silence-speech and speech-silence transitions was done using a threshold on the energy domain relative to trimmed peaks as in previous works^7-9^. The threshold was set to 65% of the 95^th^ percentile, with minimum silence length set to 20ms and minimum speech length to 30ms.

#### Statistical analysis

Descriptive results included means (standard deviations, SD), and medians (interquartile ranges, IQR). We estimated limits of agreement between microphones through the Bland-Altman method^10^ **briefly described next. In short, we calculated the difference in measurement between a pair of microphones while keeping everything else constant. For instance, we took the mean *f*o value for participant 1, reading task, recorded by the Lapel microphone, and subtracted that from the same metric (mean *f*o value) for the same participant 1, same reading task, but recorded (simultaneously) by the Reference microphone, resulting in one Lapel-Reference disagreement value for mean *f*o-participant1-reading. We them did the same process for each participant thus resulting in 18 Lapel-Reference “disagreement values” for mean *f*o value, reading task (one for each participant-task-metric).** **Unless otherwise specified, all Bland-Altman statistics below, including tests for dependency and normality, and 95% confidence intervals are calculated upon that array of “disagreement values” and not upon the original acoustic results.** Each mobile device was compared to the Reference recording device resulting in three pairwise comparisons for each acoustic measurement. **As per Bland-Altman, we then** used scatter plots of pair-wise averages by pair-wise differences in measurements to screen for outliers, measurement bias and systematic errors (i.e., **dependency of disagreement size on** mean measurement). Measurement bias was also quantified through mean differences between pairs of microphones.

We used histograms and Lilliefors tests to screen for deviations from the normal distribution in pair-wise **disagreements**. In case of non-normal distributions, we used the histogram and the sample size adjusted z-score (termed moving criterion)^11^ to screen for outliers. For our sample size, the z-score cut-off of 2.39 standard deviations corresponds to excluding the extreme 1% of differences (0.5% on each tail). We temporarily excluded the outlier. If the data then met normality assumptions, we re-included the outlier and continued with the process to determine agreement coefficients. Possible causes for the presence and degree of extreme differences are discussed later. Outliers were included in all following calculations of agreement in order to generate conservative rather than optimistic limits of agreement. When single outliers were not responsible for deviations from the normal distribution, skewness was treated by log transformation of measurements. If differences between log transformed measurements were still not normally distributed, no further analysis would be undertaken, and that dataset was reported as technically inappropriate.

Limits of agreement were defined as mean difference ± 2SD, and the coefficient of agreement as 2SD in case of non-log transformed data. For log transformed data, the equivalent antilogs of mean difference ± 2SD give a proportional result, which we multiplied by the mean measurement to inform the limits of agreement. As with non-log transformed data, the coefficient of agreement is half the difference between the upper and lower limits of agreement, and equivalent to 2 SD^10^.

The larger the sample size used for estimation of limits of agreement, the more precise the estimates. To account for precision in estimates, we calculated 95% CI for the upper and lower limits of agreement. We then expanded the concept of coefficient of agreement to include the 95% CIs and reported that as 95% CI of agreement (95% CIA, figure 2). The smaller the 95% CIA, the smaller the differences between measurements (i.e., higher agreement between microphones).
